# AROMHA Brain Health Test: A Remote Olfactory Assessment as a Screen for Cognitive Impairment

**DOI:** 10.1101/2024.08.03.24311283

**Authors:** Benoît Jobin, Colin Magdamo, Daniela Delphus, Andreas Runde, Sean Reineke, Alysa Alejandro Soto, Beyzanur Ergun, Alefiya Dhilla Albers, Mark W. Albers

## Abstract

Cost-effective, noninvasive screening methods for preclinical Alzheimer’s disease (AD) and other neurocognitive disorders remain an unmet need. The olfactory neural circuits develop AD pathological changes prior to symptom onset. To probe these vulnerable circuits, we developed the digital remote AROMHA Brain Health Test (ABHT), an at-home odor identification, discrimination, memory, and intensity assessment.

The ABHT was self-administered among cognitively normal (CN) English and Spanish speakers (n=127), participants with subjective cognitive complaints (SCC; n=34), and mild cognitive impairment (MCI; n=19). Self-administered tests took place remotely at home under unobserved (among interested CN participants) and observed modalities (CN, SCC, and MCI), as well as in-person with a research assistant present (CN, SCC, and MCI).

Olfactory performance was similar across observed and unobserved remote self-administration and between English and Spanish speakers. Odor memory, identification, and discrimination scores decreased with age, and olfactory identification and discrimination were lower in the MCI group compared to CN and SCC groups, independent of age, sex, and education.

The ABHT revealed age-related olfactory decline, and discriminated CN older adults from those with cognitive impairment. Replication of our results in other populations would support the use of the ABHT to identify and monitor individuals at risk for developing dementia.

## Introduction

Alzheimer’s disease (AD) affects over 6.9 million Americans, and this number is expected to grow to 13.9 million by 2060 with devastating economic consequences for society (>$335B / year in the US) and families (>$330B in unpaid care provided predominantly by family members)^1^. The dementia syndrome of AD is now considered an advanced stage of the disease since radiological and pathological evidence demonstrate that pathology begins to accumulate 15-20 years before the onset of memory symptoms^2–5^. At the onset of self-reported memory symptoms, neuropsychological testing is often normal – a stage termed subjective cognitive decline or subjective cognitive complaints (SCC)^6^. As the disease progresses to amnestic memory deficits revealed by psychometric testing, this stage becomes mild cognitive impairment (MCI), a stage preceding dementia where activities of daily living are not impaired yet by cognitive deficits. While most clinical trials have focused on the symptomatic stage of the disease, many investigators hypothesize that treatment during these preclinical SCC and MCI stages is likely to be more efficacious^7–9^. A cost-effective, noninvasive screen for preclinical AD performed at home would enable important research in this area by affording a means for more efficient screening, such as blood-based biomarkers and imaging, for eligibility criteria for clinical trials targeting preclinical or early-stage disease^10–12^.

The measurement of early olfactory impairment is a prime candidate as a component of an early detection assessment^13^. Many brain regions process olfactory input from primary olfactory neurons^14–16^, and these regions are damaged early in the disease – with both the olfactory bulb and entorhinal cortex among the first sites of tau pathology^17^. The amygdala and piriform cortices are also early sites of tau pathology^17,18^. In addition, the olfactory epithelium shows evidence of amyloid and tau deposition^19^. MRI studies demonstrated reduced olfactory bulb volume in AD patients and a smaller primary olfactory cortex (i.e., piriform cortex, amygdala, and entorhinal cortex) in MCI compared to cognitively unimpaired older adults^20–23^.

The hypothesis that cognitive processing of odor input may be compromised at early stages of the disease has been tested predominantly with smell identification performance, usually assessed by forced-choice measures like the University of Pennsylvania Smell Identification Test (UPSIT)^24^ or Sniffin Sticks^25^ where the four odor name choices are viewed prior to or in parallel with sniffing the odor. Smell identification performance has been associated with AD biomarkers, elevated levels of CSF and PET tau^18,26–30^ and worse performance is associated with smaller hippocampal volume in older adults^31,32^ and in patients with cognitive impairment on the AD clinical continuum^22,33–37^. Smell identification score is related to declarative memory in older adults^38^ and survives in models to predict the conversion from MCI to dementia^14,39–43^. Furthermore, smell identification scores have been shown to help predict cognitive decline in cognitively unimpaired older adults^44–49^ and the conversion to MCI^42,50,51^.

However, additional olfactory cognitive assessment tasks that probe other neural circuits vulnerable to aging and neurodegeneration could add sensitivity and specificity for olfactory screens of early damage in aging and a variety of neurodegenerative diseases, including AD, Parkinson’s^14,52^ and Traumatic Brain Injury^53^. For instance, odor memory and olfactory discrimination tasks have been associated with earlier preclinical stages of the disease^43,48,54^, and selective odor memory deficits, after correction for odor identification and odor discrimination performance, have been associated with AD biomarkers^43^. The incorporation of self-confidence within olfactory testing could additionally improve the sensitivity and specificity of olfactory testing since metacognition and self-awareness were found to be predictive of cognitive decline and biomarkers in patients with AD or MCI^55–59^.

While olfaction has been suggested as a potential screening tool for AD, logistical challenges and questions of specificity have hindered its widespread adoption^43,60,61^. To address these limitations, we developed a battery of olfactory tests. This battery includes an odor percept identification (OPID) test, where participants smell an odor, answer a question, and then choose from four provided odor names. The battery also includes a percepts of odor episodic memory (POEM) test, where participants distinguish between new odors and those presented earlier; and an odor discrimination (OD) test, where participants identify pairs of smells as either the same or different. This battery was administered using an olfactometer to deliver odors in earlier work, and we demonstrated selective odor memory loss in participants at risk of developing MCI^43^. In response to the urgency of the COVID pandemic and the respiratory transmission of the SARS-CoV2 virus, our team moved to the use of one-time use labels with embedded odors and developed an abbreviated COVID Smell Test^62^ as an early at-home screen for SARS-CoV2 infection. Subsequent testing using one-time odor labels in both English and Spanish in 30 states and Puerto Rico^63^, as well as in Argentina (manuscript in preparation), provided valuable pilot data that allowed us to adapt our olfactory battery into a bilingual at-home self-administered brain health test to screen for both nasal and cognitive deficits in processing olfactory deficits.

Here, we describe and validate this digital accessible remote olfactory-mediated health assessment: the AROMHA Brain Health Test (ABHT). We aimed (1) to validate unobserved remote self-administration of the ABHT by comparing results with observed remote self-administration and (2) to compare the results of the ABHT between cognitively healthy English and Spanish-speaking populations. We also aimed to validate the ABHT by comparing metrics between a group of clinical anosmic patients and a control group of cognitively normal individuals. Finally, we aimed to assess the feasibility of using the ABHT in a population at risk of dementia due to AD by determining whether it is sensitive to aging and cognitive decline in a cohort of cognitively normal healthy adults without cognitive complaints (CN), with subjective cognitive complaints (SCC), and with MCI.

## Results

### Design and Implementation of the AROMHA Brain Health Test

The workflow of the self-administered ABHT includes tests of odor percept identification (OPID), percepts of odor episodic memory (POEM), and odor discrimination (Fig. 1), which parallels our previous researcher-administered tests, where odors were delivered through an olfactometer^43^ or through hand-held, repeat use devices (Whispis)^64^. The ABHT leverages the remote administration aspects of our COVID Smell Test^62^ by delivering the odor stimuli using odor labels arrayed on mailable cards, by including an odor intensity measure, and by enabling self-administration by developing a web-based platform^65^ to collect responses. Additionally, the ABHT adds a meta-cognition measure embedded in the odor percept identification tasks. Participants were instructed by the web-based app to sample the odor, and then choose an odor name from a forced choice list of 4 options. They are then asked to evaluate their confidence in each odor identification decision with a scale that includes the following options: “I Guessed,” “I Narrowed Down to Three,” “I Narrowed Down to Two,” or “I Am Certain.”. This confidence metric is quantified for the OPID9 and OPID18 odor identification tests as the number answered correct among items paired with the “I am Certain”, “I Narrowed Down to Three”, and “I Narrowed Down to Two” responses (OPID9noguess, OPID18noguess scores).

**Figure 1.**
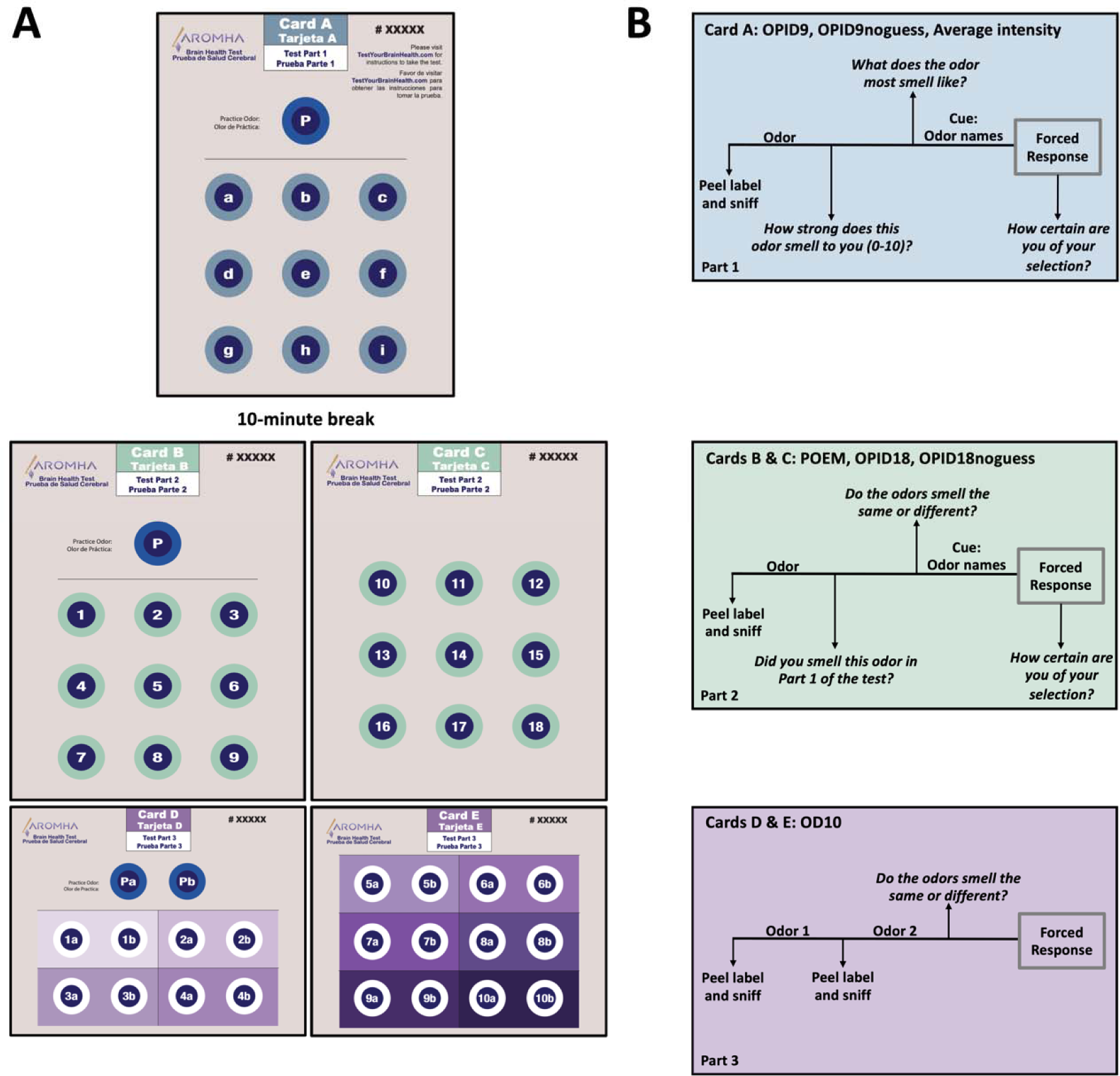
AROMHA Brain Health Test Schematic. Following online prescreening, online consent, the web-based program instructs you through the 5 bilingual (English / Spanish) cards (A). Card A is comprised of a practice odor P followed by the 9 odor labels comprising the OPID9 test. Adjacent in the blue box (B) is the workflow for these tests as directed by the testyourbrainhealth.com software to generate the OPID9, OPID9noguess, and average intensity scores. After a 10-minute break, participants are instructed to work through Cards B and C using the workflow in the green box (B) to generate the POEM, OPID18, and OPID18noguess scores. Then participants are instructed to move on to Cards D and E using the workflow in the purple box (B) to generate an OD10 odor discrimination score.

Three sets of bilingual (English / Spanish) cards, which are arrayed with odor labels, were designed to administer the OPID9 odor percept identification and odor intensity test (Part 1, Card A), the OPID18 odor percept identification and POEM memory test (Part 2, Cards B and C), and the OD10 odor discrimination test (Part 3, Cards D and E) (Figure 1). Numerous concentrations of each odor were packaged in different labels, and perceptions of odor intensity using a 10-point Likert scale that ranges from 0 (no odor) to 10 (strongest odor imaginable) were obtained from healthy college-aged participants in pilot studies. The final label-embedded odorant concentrations were selected with a mean perceived intensity of 7-7.5. To ensure that the headspace for each label was not contaminated by one or more components of the adhesive to hold the labels together, we performed gas chromatography / mass spectrometry for each odor label (Extended Data Figure 1). We did not find a common component in the headspace of all labels, which could confound olfactory performance.

We revamped the foil choices so they are more orthogonal to the target odor and more specifically evocative rather than generalized. For example, we avoided using foils describing fruit odors when the target odor was a fruit. We also incorporated foil names like “coconut” or “fresh bread” in order to include evocative odor names. We specifically avoided foils that may have a contextual association with the target odor because they are often co-presented in the real world, to reduce bias from the other foils. Finally, we expanded our set of odor names so that each odor name was only presented once within the OPID9 or OPID18 odor identification tests, either as a target odor or a foil, (Extended Data Tables 1& 2). When the 9 odors, presented in the first OPID9 odor identification and intensity test, were presented again (after a 10-minute break) in the OPID18 odor identification and POEM memory test, new sets of foils were presented with the correct name for each target odor to vary the identification experience and reduce learning carryover from OPID9 (Extended Data Tables 1& 2).

### Validation of Unobserved Remote Testing of the AROMHA Brain Health Test in Cognitively Normal Individuals

In the first phase of self-administered testing (between 5/9/23 and 8/11/23), all participants completed the ABHT in an observed setting (remote via Zoom or in person) during a scheduled appointment with the research assistant (n = 70). Participants shared their testing screen and video with the research assistant via Zoom during remote observed testing or completed the testing in person with their testing screen visible to the research assistant. In both these conditions, the research assistant observed and noted participant interactions with the software and the cards and remained on standby if there were questions or confusion. Once we felt confident that the self-administered testing workflow ran smoothly, we offered an unobserved (vs observed) self-administration option to interested cognitively normal participants in order to validate the feasibility of this administration mode and our ability to scale future data collection. When cognitively healthy participants, enrolled in our study after 8/11/23, were given the option to self-administer remotely, independent of a research assistant, 70% chose to test on their own with the option of live help over the telephone, if needed. The overall distribution of participants by each administration modality is described in Extended Data Table 3.

When comparing olfactory scores between CN observed and unobserved groups, there were no significant differences in the olfactory outcomes after Bonferroni correction. Age (*p* < .001) was significantly greater and Spanish (*p* < .001) language was significantly more common in the unobserved group, due to operational factors such as the availability of the unobserved option during phases of recruitment, scheduling, and participant preference. Test duration (*p* < .001) was also significantly greater in the unobserved group compared to the observed group, which may have biased the unobserved group to perform less well on olfactory measures. Sex and education were not different between the observed and unobserved groups (Table 1).

**Table 1.** Demographic information and olfactory function in cognitively normal participants who underwent observed and unobserved self-administration conditions of the AROMHA Brain Health Test. Values are means (SD). T-tests were performed across groups for age, education, olfactory scores, and test duration. Chi-square test was performed on sex and speaking-language proportions across groups. **Bold** indicates a statistically significant difference that remained significant after Bonferroni correction.

|  | CN Observed<br>Self-administration<br>(n = 71) | CN Unobserved<br>Self-administration<br>(n = 56) | *p* values |
| --- | --- | --- | --- |
| Age (years) | 44.89 (22.18)<br>(19 to 93 years) | 67.77 (16.60)<br>(20 to 92 years) | **< .001** |
| Sex (female %) | 66% | 64% | .82 |
| Education | 16.80 (3.86) | 16.68 (2.99) | .84 |
| Speaking language (Spanish %) | 51% | 9% | **< .001** |
| OPID9 (/9) | 5.93 (1.38) | 5.89 (1.40) | .88 |
| OPID9noguess (/9) | 5.54 (1.58) | 5.25 (1.92) | .37 |
| OPID18 (/18) | 11.85 (2.79) | 11.05 (2.57) | .10 |
| OPID18noguess (/18) | 10.83 (3.24) | 9.46 (3.57) | .03 |
| OD10 (/10) | 8.59 (1.23) | 8.36 (1.41) | .33 |
| POEM (-1 to +1) | 0.36 (0.32) | 0.33 (0.28) | .57 |
| Average intensity (/10) | 5.53 (1.83) | 5.79 (1.70) | .41 |
| Test Duration (minutes) | 31.87 (8.70) | 41.16 (16.35) | < .001 |

### Validation of the AROMHA Brain Health Test in Anosmic patients

Table 2 displays the demographics and olfactory scores of 7 patients with anosmia recruited from a smell loss clinic and CN participants. As expected, the anosmic group performed significantly worse on every olfactory metric (p <.001) in our battery, as compared to the CN group although they did not take significantly longer to complete the battery when grouped across various modalities (Extended Data Table 3). The anosmic group did not perform statistically different from chance performance on every olfactory measure, which was not the case for the CN control group.

**Table 2.** Distribution of olfactory scores of anosmic patients and CN participants on the AROMHA Brain Health Test. The first column lists the score following random selection of the answers for each test, e.g., the participant had no olfactory information to guide selection of the answers. Values are means (SD). NA = not applicable. Wilcoxon signed-rank tests were performed across groups for age, education, olfactory scores, and test duration. The chi-square test was performed for sex. **Bold** indicates a statistically significant difference that remained significant after Bonferroni correction.

|  | Score of chance performance | Anosmic (n=7) | CN (n=127) | P values |
| --- | --- | --- | --- | --- |
| Age (years) | NA | 54.14 (20.28)<br>(23 to 77 years) | 54.98 (22.88)<br>(19 to 93 years) | .91 |
| Sex (female %) | NA | 57% | 65% | .97 |
| Education (years) | NA | 16.43 (2.44) | 16.75 (3.49) | .96 |
| OPID9 (/9) | 2.25 | 2.57 (0.79) | 5.91 (1.38) | <.001 |
| OPID9noguess (/9) | 2.25 | 1.00 (1.29) | 5.41 (1.14) | <.001 |
| OPID18 (/18) | 4.5 | 5.00 (1.29) | 11.50 (2.71) | <.001 |
| OPID18noguess (/18) | 4.5 | 1.14 (1.46) | 10.23 (3.45) | <.001 |
| OD10 (/10) | 5 | 6.14 (1.57) | 8.49 (1.31) | <.001 |
| POEM (-1 to +1) | 0 | -0.02 (0.10) | 0.35 (0.30) | <.001 |
| Average intensity (/10) | NA | 1.80 (1.33) | 5.64 (1.77) | <.001 |
| Test Duration (minutes) | NA | 33.46 (11.33) | 35.96 (13.42) | .50 |

**Table 3.** Demographic information and olfactory function in cognitively normal English-speaking and Spanish-speaking participants. Values are means (SD). T-tests were performed across groups for age, education, clinical variables, olfactory scores, and test duration. Chi-square test was performed on sex and speaking-language proportions across groups. **Bold** indicates a statistically significant difference that remained significant after Bonferroni correction.

|  | CN English-speaking<br>participants<br>(n = 86) | CN Spanish-speaking<br>participants<br>(n = 41) | *p* values |
| --- | --- | --- | --- |
| Age (years) | 59.16 (23.36)<br>(19 to 93 years) | 46.20 (19.32)<br>(19 to 79 years) | **.001** |
| Sex (female %) | 63% | 71% | .38 |
| Education | 16.47 (2.84) | 17.34 (4.56) | .26 |
| OPID9 (/9) | 5.86 (1.46) | 6.02 (1.21) | .51 |
| OPID9noguess (/9) | 5.33 (1.86) | 5.59 (1.47) | .40 |
| OPID18 (/18) | 11.28 (2.92) | 11.95 (2.18) | .15 |
| OPID18noguess (/18) | 10.00 (3.62) | 10.71 (3.04) | .25 |
| OD10 (/10) | 8.42 (1.35) | 8.63 (1.22) | .37 |
| POEM (-1 to 1) | 0.31 (0.30) | 0.42 (0.30) | .06 |
| Average intensity (/10) | 5.62 (1.77) | 5.70 (1.80) | .82 |
| Test Duration (minutes) | 36.28 (14.63) | 35.30 (10.58) | .67 |

### Equivalence of the AROMHA Brain Health Test in English vs. Spanish-speaking Cognitively Normal Participants

When assessing the potential effect of language, no significant differences were found across olfactory scores between cognitively normal English-speaking and Spanish-speaking participants. Demographically only age was significantly different, as the English-speaking group was significantly older than the Spanish-speaking group (*p* = .001) (Table 3).

### Diminished Olfactory Measures in the AROMHA Brain Health Test with increasing age

When assessing the effect of age on olfaction, linear regression models of olfactory scores as a function of age of the CN participants showed that greater age was significantly associated with lower OPID9noguess (*β* = -0.02, *p* = .002), OPID18 (*β* = -0.04, *p* < .001), lower OPID18noguess (*β* = -0.06, *p* < .001), lower OD10 (*β* = -0.02, *p* < .001), and lower POEM (*β* = -0.004, *p* < .001) scores, while the association with OPID9 (*β* = -0.01, *p* = .01) and average intensity (*β* = -0.007, *p* = .28) scores did not reach significance after Bonferroni correction (Figure 2).

**Figure 2.**
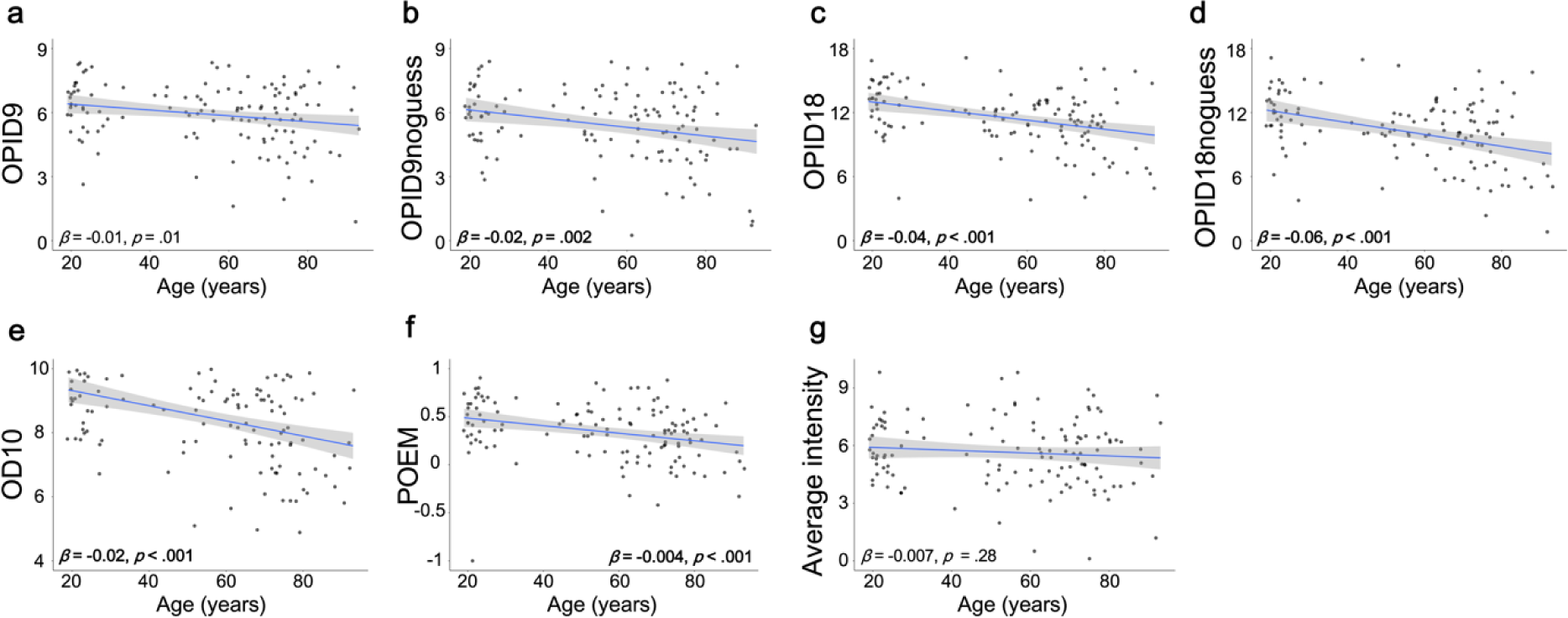
Linear regression model between olfactory scores and age across CN participants. Greater age was significantly associated with lower (b) OPID9noguess (β = -0.02, SE = .007, *p* = .002, adj. R^2^ = .06), (c) OPID18 (β *= -0.04, SE = 0.001, p < .001, adj. R*^2^ *= .12*), (d) OPID18noguess (β *= -0.06, SE = 0.03, p < .001, adj. R*^2^ *= .13*), (e) OD10 (β *= -0.02, SE = 0.005, p < .001, adj. R*^2^ *= .16*), and (f) POEM (β *= -0.004, SE = 0.001, p < .001, adj. R*^2^ *= .08*) scores, while the association with (a) OPID9 (β *= -0.01, SE = 0.005 p = .01, adj. R*^2^ *= .04*) and (f) average intensity (β *= -0.007, SE = 0.007, p = .28, adj. R*^2^ *= .001*) scores did not reach significance after Bonferroni correction for 21 comparisons (*p* < .002).

### Performance of AROMHA Brain Health Test Distinguishes Participants Aged +55 who are Cognitively Normal, with Subjective Cognitive Concerns or with Mild Cognitive Impairment

The sex and education of participants across the CN, SCC, and MCI groups were not significantly different after Bonferroni correction. Participants in the MCI group were, as expected, older than those in the CN group (p=0.01). Comparisons of each olfactory score of the ABHT for participants who are CN, have SCC, or have MCI reveal significant olfactory differences between subgroups, other than in the OD10, POEM odor memory score, and evaluations of average odor intensity (Table 4). Comparisons across cognitive groups in this +55 sample revealed no demographic differences in sex and education, or in the test duration time.

**Table 4.** Olfactory function across cognitive status among participants aged 55+. Values are means (SD). T-tests were performed across groups for age, education, clinical variables, olfactory scores, and test duration. Chi-square test was performed on sex proportion across groups. **Bold** indicates a statistically significant difference that remained significant after Bonferroni correction for 21 comparisons (*p* < .002).

|  | CN<br>(n=73) | SCC<br>(n=31) | MCI<br>(n=19) | *p* values |  |  |
| --- | --- | --- | --- | --- | --- | --- |
|  |  |  |  | CN vs.<br>SCC | CN vs.<br>MCI | SCC vs.<br>MCI |
| Age (years) (56 to 95 years) | 72.25 (9.13) | 75.48 (9.22) | 78.00 (8.07) | .11 | .01 | .32 |
| Sex (female %) | 63% | 62% | 53% | .99 | .57 | .76 |
| Education (years) | 16.42 (3.62) | 16.26 (2.50) | 16.53 (3.49) | .78 | .91 | .77 |
| OPID9 (/9) | 5.71 (1.51) | 5.19 (1.45) | 3.95 (1.43) | .10 | < .001 | .005 |
| OPID9noguess (/9) | 5.16 (1.93) | 4.29 (1.79) | 3.16 (1.77) | .03 | < .001 | .04 |
| OPID18 (/18) | 10.79 (2.74) | 10.68 (2.95) | 7.26 (2.86) | .62 | < .001 | < .001 |
| OPID18noguess (/18) | 9.4 (3.47) | 8.03 (4.02) | 5.16 (3.35) | .11 | < .001 | .009 |
| OD10 (/10) | 8.12 (1.35) | 7.42 (1.52) | 6.95 (1.96) | .03 | .02 | .38 |
| POEM (-1 to +1) | 0.27 (0.29) | 0.24 (0.25) | 0.22 (0.20) | .61 | .37 | .72 |
| Average intensity (/10) | 5.55 (1.85) | 4.93 (1.34) | 4.71 (2.30) | .06 | .15 | .71 |
| Test Duration (minutes) | 40.10 (14.67) | 40.49 (14.89) | 40.69 (11.01) | .66 | .85 | .83 |

When assessing the effect of cognitive status group on olfactory test components, ANCOVAs revealed a significant effect on OPID9, OPID9noguess, OPID18, OPID18noguess, and OD10 scores (Table 5). No effects of cognitive status group were found for the POEM or average intensity scores. After Bonferroni correction for 21 comparisons (p < .002), post-hoc pairwise comparisons revealed significantly lower scores in the MCI group compared to CN older adults for the OPID9, OPID18, OPID18noguess, OD10 and to the SCC group for the OPID18 and OPID9 scores.

**Table 5.**
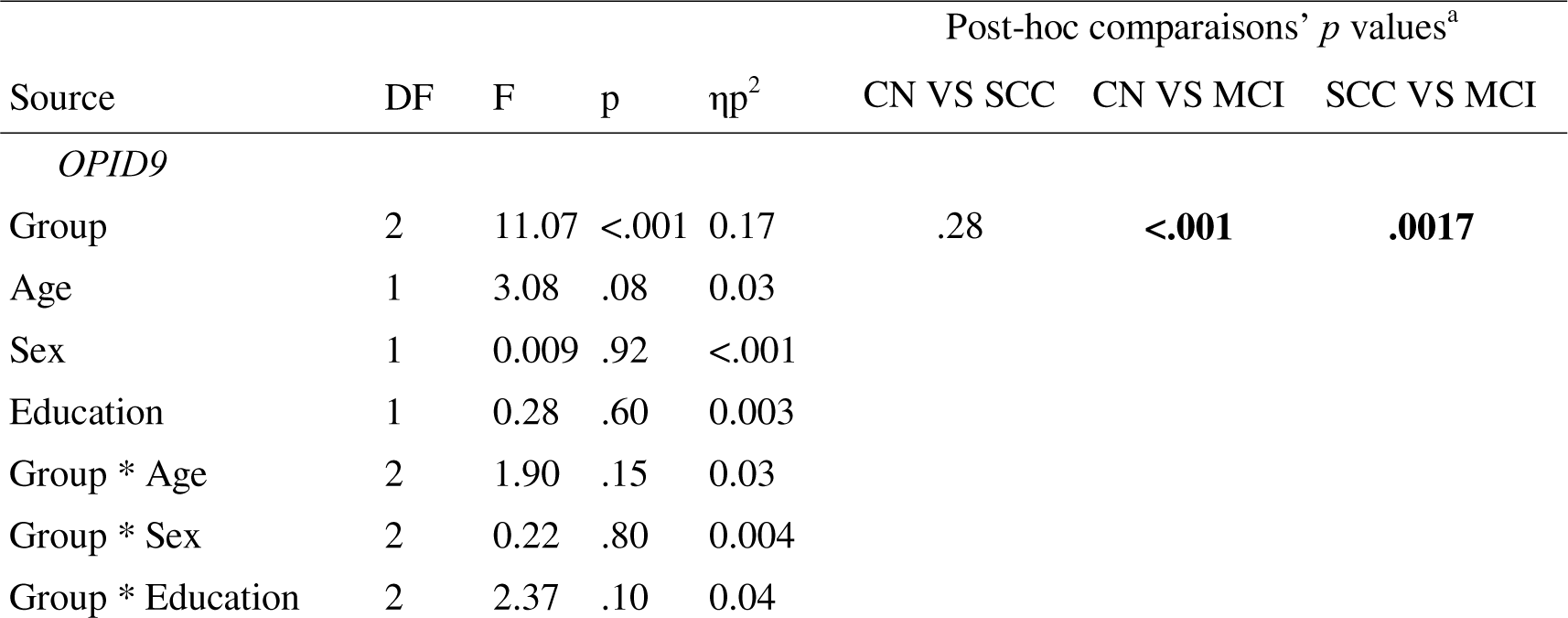
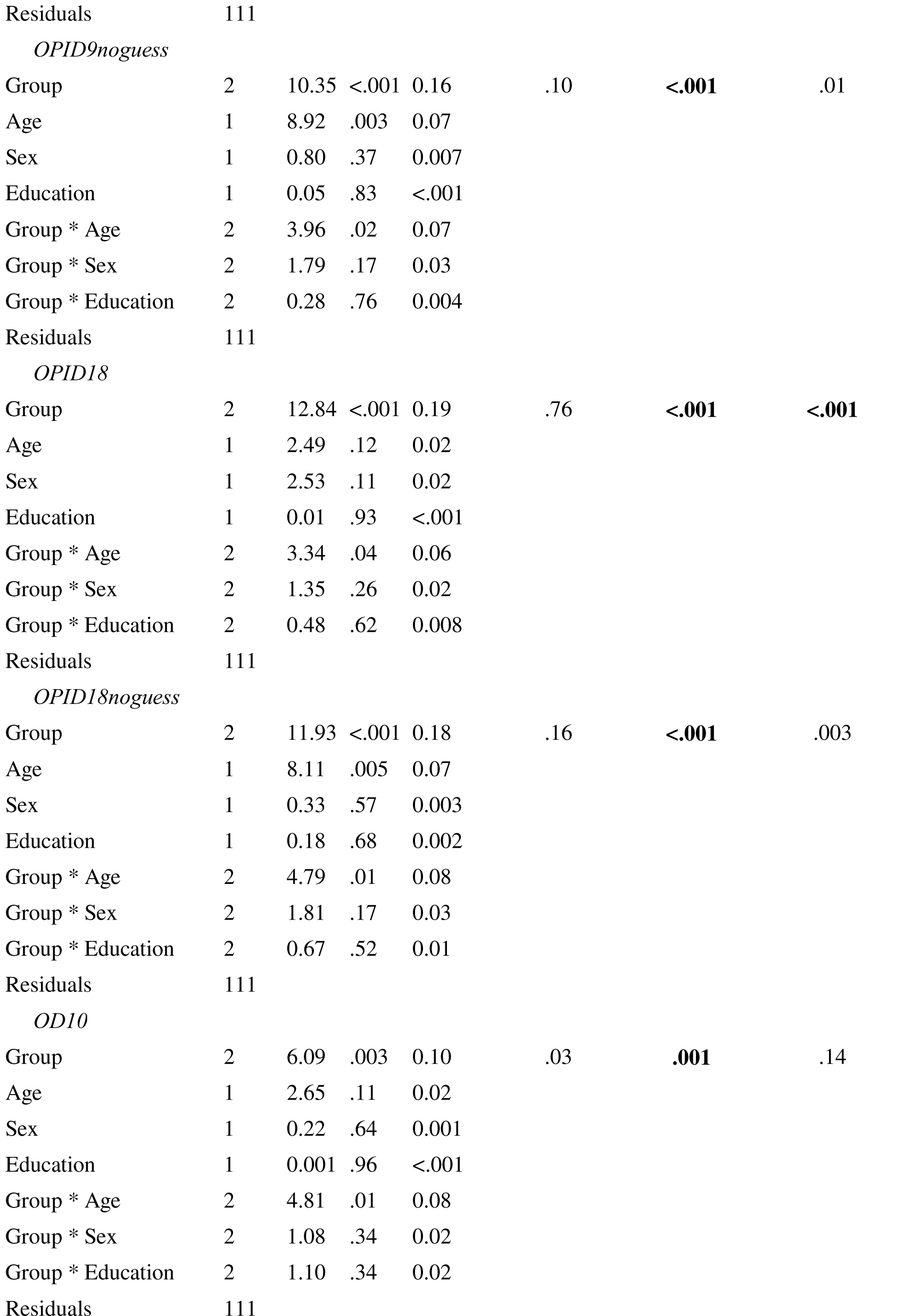
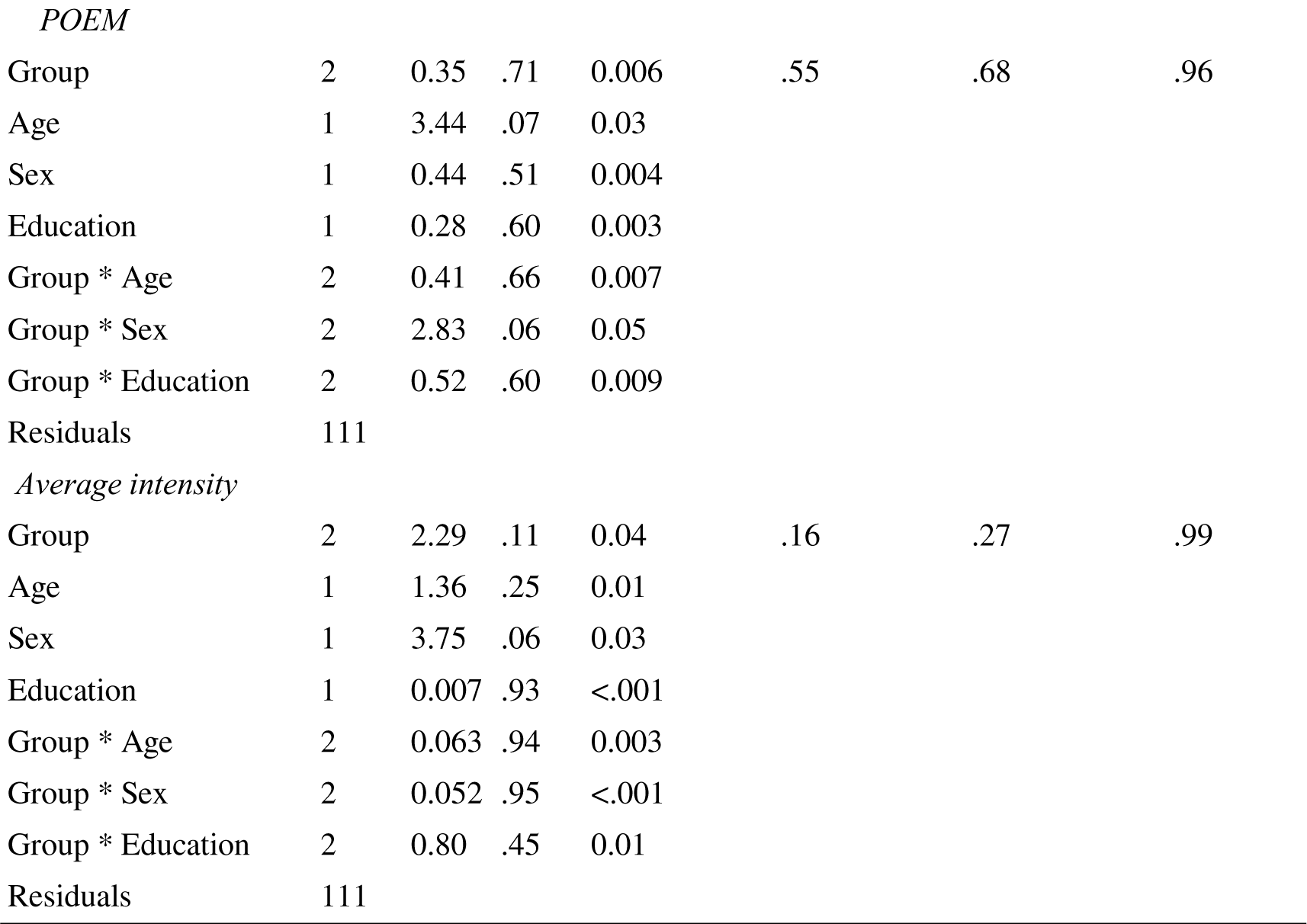
ANCOVA models comparing different olfactory scores across groups. DF = Degrees of Freedom, F = F-statistic, p = p-value, ηp^2^ = partial Eta squared. CI = Confidence interval. **Bold** represents post-hoc *p* values that remained significant after Bonferroni correction for 21 comparisons (*p* < .002). ^a^ *p* values from pairwise post-hoc comparisons adjusted for age, sex, and education.

Interaction effects were found with age for the OPID9noguess, OPID18 no guess scores, and OD10, however they did not remain significant after Bonferroni correction for 7 comparisons (p < .007). Other olfactory components did not interact with age (*p* > .05). No interaction effect with sex or education were found with any olfactory scores (*p* > .05) (Table 4).

## Discussion

In this study, we tested and validated a self-administered olfactory test battery on cognitively healthy English and Spanish-speaking cohort in the home setting, under observed and unobserved self-administered conditions. We began with observed self-administered testing of participants who were cognitively normal, had expressed subjective cognitive concerns (SCC), and had a diagnosis of Mild Cognitive Impairment (MCI). Both participants with SCC and MCI are at risk of developing Alzheimer’s disease dementia^66,67^. The majority of this observed testing was completed with the convenience of a remote in-home setting, using Zoom to share the testing screen and a video of the participant with a research assistant observing the workflow, noting challenges, and being available if questions or concerns arose. Some of the participants chose to self-administer the test without the Zoom interface and with a research assistant in the room for questions because they preferred this or found it more convenient. After about three months of observed and largely remote self-administered testing across the spectrum of cognitive impairment (from CN to MCI), we shifted to a completely unobserved remote testing option for any cognitively normal participants who felt comfortable engaging with the test entirely on their own. When given the option to self-administer the test remotely, independent of a research assistant, the majority of cognitively healthy participants, representing a wide age spectrum ranging from 20 to 92 years old, chose unobserved self-administration.

We found equivalent olfactory performance when comparing observed and unobserved self-administration of the ABHT among CN participants in unadjusted analyses, other than shorter test duration and a younger population on average in the observed setting. We also validated the test on an anosmic subsample, demonstrating that anosmic patients performed as expected at chance level for each olfactory test. When comparing the olfactory battery scores between languages, no differences were found regarding olfactory subtest scores between CN English and Spanish-speaking populations in unadjusted analyses. We also found that lower odor percept identification (OPID9 and OPID18noguess and OPID18 scores), odor memory (POEM) and odor discrimination (OD10) scores were negatively associated with age. Finally, by comparing olfactory scores between participants aged 55+ with CN, SCC, and MCI, we found that the means of all odor percept identification scores (OPID9, OPID9noguess, OPID18, OPID18noguess) and the odor discrimination score were associated with cognitive decline, i.e., lower scores in the MCI subgroup relative to the older CN subgroup. The negative relationships observed in our analyses between identification measures adjusted for metacognition (OPID9noguess and OPID18noguess) and increasing age and cognitive impairment are intriguing, given prior work on the general prevalence of olfactory overconfidence in relation to the emotionality of odors and eventual identification accuracy^68,69^. Further work on these new “noguess” measures is needed in relation to AD biomarkers and longitudinal outcomes in order to better understand this finding.

By leveraging our experience during the COVID pandemic, we were able to successfully adapt and evolve our previous brain health test for remote at home self-administration in our target populations of interest. This included protocols for online screening for eligibility, online consenting, and an optimized remote user interface to guide participants through various olfactory tasks. We also modified odor delivery by using odor labels on mailable cards. Unlike our COVID test^62,63^, the ABHT is particularly targeted for participants aged 55+, a population generally less familiar with fully online test administration. We enrolled participants as old as 88 in the CN and SCC groups and 95 in the MCI group, who were able to successfully self-administer the test and enter responses on the web-based platform. The remote administration paradigm afforded participation from 21 different US states and Puerto Rico through online recruitment via an MGH and clinicaltrials.gov websites. The robust engagement of older participants in entirely independent, unobserved testing—yielding olfactory performance outcomes statistically indistinguishable from those obtained in observed settings—constitutes a significant finding. This challenges the prevailing assumption that older individuals are either unable or unwilling to effectively engage in self-administered remote screening methods such as ours. This screen was designed to address the unmet need of early detection of biomarkers in participants who are CN and those who have SCC but may not meet testing thresholds for MCI.

We recruited patients with MCI to validate the efficacy of this screen to detect differences in our olfactory tests under conditions of more apparent cognitive impairment, as we and others previously demonstrated^40,43,60,70^. Each olfactory identification score from the ABHT, including the 9-item sub scores (i.e., OPID9, OPID9 no guess), was lower in the MCI group compared to CN aged +55 groups. This result indicates that the ABHT could be used as a marker of cognitive decline in older adults and replicates the findings of two meta-analyses that demonstrated a specific pattern of olfactory impairment targeting more severely olfactory identification in patients with Alzheimer’s disease dementia^52^ and MCI^70^. This early decline in odor identification likely reflects damage in specific limbic and medial temporal lobe olfactory areas involved in olfactory identification in the earliest stages of AD (i.e., piriform cortex, amygdala, entorhinal cortex, hippocampus)^20,21,31–37,71,72^. At the cognitive level, olfactory identification tasks are associated with declarative memory^38,73^ and are predictive of cognitive decline in CN older adults^44–49^ and of the conversion to MCI^42,50,51^. Future studies should aim to replicate these findings and assess the predictive value of the ABHT and odor percept identification sub-scores on AD biomarkers and longitudinal cognitive decline in CN older adults.

Olfactory discrimination (OD10) was also lower in the MCI group compared to CN aged 55+ group. This result replicates and is aligned with the results of previous studies showing a lower olfactory discrimination performance in MCI^54^ and predictive value of olfactory discrimination in further cognitive decline^48^. Unlike the basic detection of odors, both identification and discrimination of odors require high-level cognitive functions such as working memory and decision making, which could explain our results^54^. The hippocampal network is also associated with olfactory discrimination^74^, early damages to this structure in AD could explain these findings. Future studies should investigate the relationship between hippocampal damage and olfactory discrimination tasks in participants at risk of AD using volumetric magnetic resonance imaging and tau PET^18^.

Objective measurements of olfactory identification, discrimination, and memory were negatively associated with age in the asymptomatic CN sample. These results are not surprising and replicate the robust literature on the effect of normal aging on olfactory function. Indeed, according to a meta-analysis including 175,073 participants (18-101 years), olfactory impairment would be prevalent at 34.5% in studies with a mean age above 55 years, while the prevalence would be at 7.5% in studies with a mean age below 55 years^75^. The decline starts in the fifth decade of life^76^ and is general across different olfactory capacities such as olfactory detection, discrimination, identification^77^, and memory^78,79^. Our results indicate that perceived olfactory average intensity was not related to age. While some studies using magnitude estimation procedures to assess the perceived intensity of odors across different concentrations found no significant age-related differences^80,81^, another study found that older individuals perceived increases in menthol concentration as less intense than younger individuals^82^. These mixed results suggest that while perceived odor intensity may not always show a clear age-related decline, other aspects of olfactory function, such as identification, discrimination, and memory, are consistently negatively impacted by aging.

Having validated remote unobserved self-administration in CN people and demonstrated that remote observed self-administration of the ABHT is feasible in an MCI population (95% of MCI participants completed testing in this mode), and by finding expected differences in olfactory identification scores as shown in previous studies, we are poised to begin studies in deeply phenotyped populations to quantify the predictive value of ABHT outcomes on biomarkers of neurodegenerative disease, including Alzheimer’s, Lewy Body disease, and concussive and non-concussive head trauma. These studies can incorporate both English and Spanish-speaking participants, as we did not find any differences in olfactory scores among the CN English and Spanish-speaking participants. Different olfactory-behavioral profiles are hypothesized to emerge depending on disease neuropathology, as different brain areas and networks of the central olfactory system are associated with different olfactory tasks^83–85^. While previous studies showed that MCI and AD predominantly affect olfactory identification^52,70^, the olfactory bulb is affected at the earliest stage of Parkinson’s disease resulting in a general olfactory impairment with reduced olfactory detection performance and leads to a decrease in various olfactory tasks (e.g., discrimination of odors, identification of odors)^52,86^. Other conditions such as Lewy body dementia^87^, frontotemporal dementia^88^, and exposure to head impacts and traumatic brain injury^89–91^ can also cause olfactory impairments.

When combined with other digital biomarkers, a mobile self-administrated and diverse smell assessment like ours could accelerate screening for neurodegenerative diseases in asymptomatic or newly symptomatic individuals who would benefit from more definitive subsequent tests^92,93^, such as blood-based, image-based, or cerebrospinal fluid (CSF)-based diagnostics, especially in individuals presenting additional risk factors for dementia such as subjective cognitive decline^66^, depression^94^, and genetic risks factors such as the APOE-4 allele^95^. Home-based tests also have the potential to enhance the involvement of underrepresented groups in research settings^96^ and to save time and cost of transportation, to save costs for the healthcare system, and to increase patient satisfaction^97,98^.

This study has certain limitations. The main limit is the restricted availability of neuropsychological testing for a proportion of the participants (unverified vs verified) in the sample. Additionally, the results of this study correlate with a robust body of literature, and demonstrate its utility in real-world settings. Future longitudinal studies will assess the predictive value of the ABHT on longitudinal cognitive decline and highlight the utility of this olfactory battery in mapping correlations between patterns olfactory decline and the progression of various neurodegenerative diseases. While not a limitation in this preliminary validation of the AROMHA Brain Health Test, biomarker availability will be an essential component in future studies that assess the predictive power that this noninvasive preliminary screen may hold.

## Conclusion

The ABHT is a novel remote olfactory battery that exhibited similar performance across observed and unobserved self-administration among CN participants as well as among English and Spanish-speaking CN participants, while anosmic patients performed at chance level as expected. Odor percept identification, discrimination, and memory subtests were sensitive to the aging effect on the olfactory system. Each olfactory identification subtest, including the short 9-item version, and the olfactory discrimination subtest showed lower performance in the MCI group, mirroring results in the literature. These results suggest that the ABHT could be used in clinical research settings in different languages to explore the utility of olfactory biomarkers to predict the presence of blood-based, image-based, or cerebrospinal fluid (CSF)-based biomarkers of neurodegenerative disease and longitudinal development of clinical symptoms.

## Methods

### Development and Quality Control of the AROMHA Brain Health Test

The ABHT was updated for remote at-home self-administration of previously developed Odor Percept Identification (OPID), Percepts of Odor Episodic Memory (POEM), and Odor Discrimination (OD) subtests^43^. All pre-screening, informed consent, and administration of the test occurs online through a web-based interface (testyourbrainhealth.com). This updated version of the test consisted of five different 8.5 “ x 11” single-use cards that were packaged in one envelope and mailed to the participant’s home. We expanded the manufacturing of odor labels from the three odors utilized in the COVID smell test^62^ to include additional 15 odors. Odor labels were manufactured by MFR Samplings using Living Library^TM^ odors purchased from International Flavors and Fragrances (IFF)^62^. Odors were presented to participants in a peel-and-sniff manner and contained proprietary naturalistic odors from the Living Library developed by IFF (https://www.iff.com/). Gas chromatography/mass spectrometry were conducted at the Mass Spectrometry core at the Bauer Laboratory in the Harvard Chemistry Department. Briefly, each odor label was completely opened in a stoppered 15 ml conical tube and allowed to reach equilibrium for 1 minute at room temperature. Then a Hamilton Syringe was used to inject a representative sample of the headspace into the GC/MS instrument. The peaks were normalized to 2-methyl-3-heptanone equivalents, and analyzed for common set of peaks that might represent a common contaminant from the adhesive. All samples were run the same day to eliminate batch effects (Extended Data Figure 1).

Participant responses to all components of the ABHT were collected on a web-based application at testyourbrainhealth.com designed for independent self-administration of the survey questions and the olfactory battery (Extended Data Figure 2). The data is stored on a HIPPA-compliant AWS server. The prescreening module and the informed consent module were developed on a RedCAP platform at the Massachusetts General Hospital. All protected health information was kept on RedCAP platform. Participants had the option to call a research assistant for live help in English or Spanish at any time during remote testing. The web-based AHBT application directed participants to a RedCap secure e-consent project to collect identifiers. Once consented, participants were sent back to the AROMHA, Inc. app to walk through all three parts of the battery and collect olfactory information associated with their card ID. The app was designed to lead participants through every stage of testing, including directions on how to peel odor labels and sample odors as well as respond to questions regarding odor intensity, odor identification & naming confidence, odor memory, and odor discrimination. The application has the ability to run in an English or Spanish language mode, based on participant preference. During testing, it collects participant responses for all aspects of the olfactory battery in addition to the timing of those inputs. The app generates summary and item-specific data on these metrics that can be downloaded by researchers for analysis and joined offline to demographic information collected in RedCap following the e-consent process. These results are not shared with participants.

#### Part 1: Odor Percept Identification Test (OPID9)

Participants first completed the OPID9, which involved identifying nine distinct odors: menthol, clove, leather, strawberry, lilac, pineapple, smoke, soap, and grape. These odors were selected for their predictive value in identifying the conversion to Alzheimer’s disease (AD) in patients with mild cognitive impairment (MCI)^39^. After each odor presentation, participants rated the intensity on a Likert scale from 0 to 10. Subsequently, they were presented with four odor names, asked to choose the label that best represented the odor they sampled, and asked to rate their confidence in their identification choice using the following scale: “I Guessed,” “I Narrowed Down to Three,” “I Narrowed Down to Two,” or “I Am Certain.” (Extended Data Figure 2B). This process was repeated for all nine odors. Following the completion of the OPID9 test, there was a 10-minute delay during which participants answered demographic, medical, nasal, and memory-related questions.

#### Part 2: Percepts of Odor Episodic Memory (POEM) / OPID18

After the 10-minute break, participants completed the POEM/OPID18 tests. These tests included the nine odors from Part 1 and nine additional odors: coffee, peach, chocolate, orange, dirt, banana, lemon, bubble gum, and rose. The odors were presented in a stereotyped random order that was held consistent across all participants. For each odor, participants first indicated whether the odor sampled was presented in Part 1 (yes/no), OPID9. As in the earlier odor identification test, they then selected the odor name most representative of the odor from four choices and rated their confidence in their selection.

#### Part 3: Odor Discrimination (OD10)

In Part 3, participants were presented with 10 pairs of odors, all of which were previously presented in Parts 1 and 2. They were asked to determine whether the paired odors were the same or different (yes/no).

The POEM index was calculated as the difference between the proportion of correct and incorrect recognitions, with scores ranging from -1 to 1. OPID9 and OPID18 scores were calculated as the total number of correctly identified odors, with maximum scores of 9 and 18, respectively. The OD10 score was the total number of correctly discriminated odor pairs, with a maximum score of 10. The average intensity score was derived from the mean intensity ratings of the nine odors from Part 1 on the Likert scale. OPID9noguess and OPID18noguess scores were calculated as the total number of odors identified correctly where the participant did not select “I Guessed” for the confidence question immediately following identification.

After verifying participants’ comfort with using the web-based application in conjunction with the AROMHA Brain Health Test’s smell cards through the use of research assistant observation, cognitively healthy participants were given the option to complete the test with or without the observation of a research assistant. Verified cognitively impaired participants were not given this option and were only able to self-administer the smell test remotely or in person under the observation of a research assistant.

#### Participants

Since May 2023, 127 CN, 34 SCC, and 19 MCI participants were recruited across 21 states in the United States, and from Puerto Rico, from the Longitudinal Cohort at the Massachusetts Alzheimer’s Disease Research Center (ADRC), through an internet posting on the Massachusetts General Hospital research site, and through clinicaltrials.gov (NCT05881239). Anosmic participants were recruited from the Smell Clinic of Dr. Mark Albers at Massachusetts General Hospital.

The cognitive status classification was verified for subjects recruited from the ADRC Longitudinal Cohort (n = 59), whereas participants recruited from the internet were classified cognitively based on self-reported cognitive complaints or medical diagnoses affecting cognitive function (n = 118). All participants underwent informed consent before participation. The research protocol was conducted in accordance with the Declaration of Helsinki and approved by the Institutional Review Board of Partners Health.

Unverified CN (n = 99) were recruited from the internet and were aged 18 years old and over and reported no cognitive complaints or medical diagnoses affecting cognitive function. From the unverified CN participants, we created a subgroup of unverified CN aged 55+ (n = 45). Verified CN aged 55+ recruited from the ADRC (n = 28), had a Montreal Cognitive Assessment (MoCA) score within the normal range adjusted for age, sex, and education level, a Clinical Dementia Rating (CDR) global score of 0, and a performance within the normal range on the Logical Memory II subscale delayed paragraph recall (LM-IIa) of the Wechsler Memory Scale-Revised (WMS-R) (≥16 years of education: ≥9; 8-15 years: ≥5; 0-7 years: ≥3). In total, we recruited 127 CN participants, 73 of them were aged 55+.

Unverified Subjective Cognitive Concern (SCC) participants (n = 13) were aged 55+ and reported SCC based on the following 3 questions: 1) Have you experienced a change in your memory in the last 1-3 years? 2) Has this been a persistent change over the last 6 months? and 3) Are you concerned about this change? Responses were on a Likert scale: “Not at all,” “Slightly,” “Moderately,” “Considerably,” and “Extremely.” If participants endorse “Slightly” or worse to all 3 questions, they will be categorized as SCC. Verified SCC participants (n = 18) were assigned using the same criteria as verified CN older adults, as well as reporting significant subjective memory concerns based on the three questions presented above. Three participants under 55 reported SCC and were removed from the analyses. In total, we included 31 participants with SCC.

Unverified MCI participants (n = 4) were aged 55+ and reported a clinical diagnosis of MCI by a certified physician. Verified MCI participants (n = 15) were aged 55+, met the National Institute on Aging-Alzheimer’s Association (NIA-AA) diagnostic criteria for MCI^99^, with a performance below an education-adjusted cut-off score on the WMS-R LM-IIa (≥16 years: ≤8; 8-15 years: ≤4; 0-7 years: ≤2), or had a MoCA score indicating MCI adjusted for age, sex, and education level, had a CDR global score from 0.5 to 1 (with memory box score of 0.5 or 1), had preserved IADL (determined by a clinician), and were not demented. In total, we recruited 19 participants with MCI.

We performed Wilcoxon rank sum/Mann Whitney tests to compare the median performance on each olfactory measure between verified and unverified participants in each subgroup (CN, SCC, and MCI). Since, after adjustment for multiple comparisons, no difference was significant at a threshold of 0.05 we combined the verified and unverified participants within a subgroup for the purposes of these analyses. Clinical data comparing the verified participants in each group are presented and compared in Extended Data Table 4.

### Statistical Analysis

All analyses were conducted in R using *Base R, effectsize, emmeans, and rstatix* packages. Student T-tests were used to compare demographics and olfactory variables between observed and unobserved conditions and between English and Spanish-speaking groups, while the Wilcoxon signed-rank test was used to compare anosmic patients with CN participants. To assess the effect of age on olfaction, we conducted linear regression models with age and olfactory scores. We performed this analysis for each olfactory subtests (OPID9, OPID9noguess, OPID18, OPID18noguess, OD10, POEM, and average intensity). To assess the effect of cognitive status on olfaction, ANCOVAs were performed to compare olfactory functioning (OPID9, OPID9noguess, OPID18, OPID18noguess, OD10, POEM, and average intensity) among older adults without cognitive impairment, participants with SCC, and with MCI, including age, sex, and education as covariates. For all the analyses, we set the alpha value at 0.05 and used Bonferroni correction for multiple comparisons.

## Acknowledgements

We thank the participants and their families for their dedication to the study. We thank Dr. Jennifer Wang and Dr. Sunia Traeger from the Mass Spectrometry core at the Bauer Laboratory in the Harvard Chemistry Department for the GC/MS analysis, Jay Lockwood for software development, Dr. Matthias Tabert at IFF for advice about odors, Daniel Tater for software advice, Pablo Ripoli and Juan Manual Arias at MFR Samplings for advice about odor labels, Emily Rusk for input about quality controls and regulatory advice, Patrizia Vannini and Gad Marshall for discussions about metacognition in smell testing, Judy Johanson, Teresa Gomez-Isla M.D., Ph.D., and Brad Hyman, M.D., Ph.D. for their assistance with recruiting ADRC subjects.

BJ is supported by scholarships from the Canadian Institutes of Health Research, the Fonds de Recherche Québec Santé, the Quebec Bio-Imaging Network, the Université du Québec à Trois-Rivières, and MITACS. The study was funded by NIH R41AG062130 and NIHR42AG 062130 (awarded to S.R.), and U01DC019579 (awarded to M.W.A.).

## Author Contributions

S.R., A.D.A., and M.W.A. conceptualized and designed the study. B.J., C.M., D.D., A.R., A.A.S., B.E., acquired and analyzed data. B.J., C.M., A.R., A.D.A., and M.W.A. drafted text and prepared figures. All authors revised content.

## Competing interests

S.R. and M.W.A. are co-founders and own shares in Aromha, Inc. C.M. and A.D.A. are consultants for Aromha, Inc. M.W.A. and A.D.A.’s participation in this research was reviewed by the MGB Office of Industrial Interactions. M.W.A is a consultant for Sudo Bioscience Limited, Transposon Therapeutics, and has received in kind donations from IFF and from Eli Lilly. He has received speaking fees from Biohaven and Incyte.

## Data availability statement

The dataset generated and analyzed for the current study are available to academic researchers with a Data Use Agreement with MGH and Aromha, Inc.

## Notes

### Clinical Trial

NCT05881239

### Clinical Protocols

https://clinicaltrials.gov/study/NCT05881239?term=NCT05881239&rank=1

### Author Declarations

The research protocol was conducted in accordance with the Declaration of Helsinki and approved by the Institutional Review Board of Partners Health.

