## Extended Data Figures and Tables for "AROMHA Brain Health Test: A Remote Olfactory Assessment as a Screen for Cognitive Impairment"

Extended Data Table 1. Odors and four choices presented to participants for the OPID9 test.

| Trial | Odor Name 1 | Odor Name 2 | Odor Name 3 | Odor Name 4 |
| --- | --- | --- | --- | --- |
| Practice trial | cherry  (cereza) | glue (pegamento) | **grass**  **(césped)** | tomato sauce  (salsa de tomate) |
| 1 | lime  (lima) | soy sauce  (salsa de soya) | coconut  (coco) | **menthol**  **(mentol)** |
| 2 | barbecue  (parrilla) | **clove**  **(clavo)** | popcorn  (palomitas de maíz) | olive  (aceituna) |
| 3 | rosemary  (romero) | **leather**  **(cuero)** | onion  (cebolla) | rose  (rosa) |
| 4 | **strawberry**  **(fresa)** | cedar wood (madera de cedro) | chocolate  (chocolate) | baby powder  (talco para bebés) |
| 5 | eggs  (huevo) | basil  (albahaca) | garlic  (ajo) | **lilac**  **(lila (flor))** |
| 6 | **pineapple**  **(piña)** | chlorine  (cloro) | sunblock  (bloqueador solar) | cotton candy  (algodón de dulce) |
| 7 | chili pepper  (ají picante) | pine tree  (pino) | jasmine  (jazmín) | **Smoke**  **(humo)** |
| 8 | new car smell  (olor a carro nuevo) | fresh bread  (pan fresco) | **soap**  **(jabón)** | mango  (mango) |
| 9 | barn animal  (animal de granja) | wet paint (pintura húmeda) | **grape**  **(uva)** | apple  (manzana) |

Note: **Bold** indicates the targeted and presented odor.

Extended Data Table 2. Odors and four choices presented to participants for the OPID18 test.

| Trial | Odor Name 1 | Odor Name 2 | Odor Name 3 | Odor Name 4 |
| --- | --- | --- | --- | --- |
| Practice trial | cherry  (cereza) | glue  (pegamento) | **grass**  **(césped)** | tomato sauce  (salsa de tomate) |
| 1 | **menthol**  **(mentol)** | barbecue  (parrilla) | ginger  (jengibre) | dry-erase marker  (marcador de borrado en seco) |
| 2 | new car smell  (olor a carro nuevo) | pine tree  (pino) | fresh bread  (pan fresco) | **coffee mocha**  **(café moca)** |
| 3 | eggs  (huevo) | lavender  (lavanda) | raw fish  (pescado crudo) | **dirt**  **(tierra)** |
| 4 | rubbing alcohol (alcohol desinfectante) | **chocolate**  **(chocolate)** | mango  (mango) | wet paint  (pintura húmeda) |
| 5 | honey  (miel) | lime  (lima) | soy sauce  (salsa de soya) | **clove**  **(clavo)** |
| 6 | cheese  (queso) | cilantro  (cilantro) | onion  (cebolla) | **orange**  **(naranja)** |
| 7 | cucumber  (pepino) | popcorn  (palomitas de maíz) | **grape**  **(uva)** | basil  (albahaca) |
| 8 | barn animal  (animal de granja) | roast chicken  (pollo asado) | cinnamon  (canela) | **lilac**  **(lila (flor))** |
| 9 | **bubble gum**  **(goma de mascar)** | fresh linen  (lino fresco) | garlic  (ajo) | cedar wood  (madera de cedro) |
| 10 | cotton candy  (algodón de azúcar) | mulch (viruta/mantillo) | black pepper (pimienta negra) | **leather**  **(cuero)** |
| 11 | ocean breeze  (brisa del océano) | tobacco  (tabaco) | cola  (coca cola) | **banana**  **(banana)** |
| 12 | vanilla  (vainilla) | jasmine  (jazmín) | **smoke**  **(humo)** | baby powder  (talco para bebés) |
| 13 | **soap**  **(jabón)** | pumpkin  (calabaza) | coconut  (coco) | chlorine  (cloro) |
| 14 | peppermint  (menta) | **rose**  **(rosa)** | watermelon (melón/sandía) | chili pepper  (ají picante) |
| 15 | licorice  (regaliz (dulce)) | bleach (blanqueador/cloro) | pizza  (pizza) | **lemon**  **(limón)** |
| 16 | olive  (aceituna) | caramel  (caramelo) | **strawberry**  **(fresa)** | sunblock  (bloqueador solar) |
| 17 | apple  (manzana) | gasoline  (gasolina) | vinegar  (vinagre) | **pineapple**  **(piña)** |
| 18 | **peach (durazno/melocotón)** | brown sugar  (azúcar morena) | tar  (brea/alquitrán) | rosemary  (romero) |

Note: **Bold** indicates the targeted and presented odor.

Extended Data Table 3. Distribution of Participants by Administration Modality

|  | Anosmic (n=7) | CN  (n=127) | SCC*  (n=34) | MCI  (n=19) | Total  (n=187) |
| --- | --- | --- | --- | --- | --- |
| Self-Administered | 7 (100%) | 127 (100%) | 34 (100%) | 19 (100%) | 187 (100%) |
| Remote | 6 (86%) | 118 (93%) | 30 (88%) | 18 (95%) | 172 (92%) |
| In-person | 1 (14%) | 9 (7%) | 4 (12%) | 1 (5%) | 15 (8%) |
| Remote Observed | 1 (14%) | 62 (49%) | 15 (44%) | 16 (84%) | 94 (50%) |
| Remote Unobserved | 5 (71%) | 56 (44%) | 15 (44%) | 2 (11%) | 78 (42%) |
| In person Observed | 1 (14%) | 9 (7%) | 4 (12%) | 1 (5%) | 15 (8%) |

* Three participants with SCC were removed from the main analyses because they were less than 55 years old.

Extended Data Table 4. Clinical values and olfactory function across cognitive status in verified older adult participants.

|  |  |  |  |  | *p* values |  |
| --- | --- | --- | --- | --- | --- | --- |
|  | Verified CN older adults (n=28) | Verified SCC  (n=18) | Verified MCI  (n=13) | CN vs. SCC | CN vs. MCI | SCC vs. MCI |
| Age (years) | 72.75 (10.96) | 78.17 (9.31) | 77.69 (5.31) | .08 | .06 | .86 |
| Sex (female %) | 61% | 56% | 54% | .97 | .94 | .99 |
| Education (years) | 15.71 (4.57) | 16.11 (2.56) | 16.08 (3.04) | .71 | .77 | .97 |
| Clinical Dementia Rating (CDR) | 0.00 (0.00) | 0.42 (0.19) | 0.54 (0.14) | **< .001** | **< .001** | .049 |
| MoCA (z-score adjusted for age, sex, and education) | 0.29 (0.84) | -0.24 (1.17) | -0.84 (1.06) | .11 | **.003** | .15 |
| OPID9 (/9) | 6.07 (1.21) | 5.11 (1.64) | 3.00 (1.78) | .04 | **< .001** | **.**049 |
| OPID9noguess (/9) | 5.29 (3.67) | 4.29 (1.79) | 3.16 (1.77) | .008 | **.001** | .32 |
| OPID18 (/18) | 11.25 (2.74) | 10.00 (3.09) | 7.31 (2.95) | .08 | **< .001** | .04 |
| OPID18noguess (/18) | 9.46 (3.82) | 7.00 (3.79) | 4.77 (3.32) | .04 | **< .001** | .09 |
| OD10 (/10) | 8.29 (1.49) | 7.39 (1.69) | 7.15 (2.12) | .07 | .10 | .74 |
| POEM (-1 to +1) | 0.24 (0.27) | 0.23 (0.25) | 0.25 (0.2) | .96 | .83 | .80 |
| Average intensity (/10)  Test Duration (minutes) | 5.29 (2.11)  41.99 (20.88) | 4.71 (1.40)  42.26 (15.28) | 4.78 (2.30)  39.91 (8.64) | .27  .96 | .50  .65 | .93  .59 |

Note: Values are means (SD). T-tests were performed across groups for age, education, clinical variables, olfactory scores, and test duration. Chi-square test was performed on sex proportion across groups. **Bold** indicates a statistically significant difference that remained significant after Bonferroni correction for 21 comparisons (*p* < .002).

Extended Data Figure 1. Gas Chromatograph / Mass Spectrometry for the headspace of each odor. All samples were run on the same day. No common peaks were seen between the headspace.

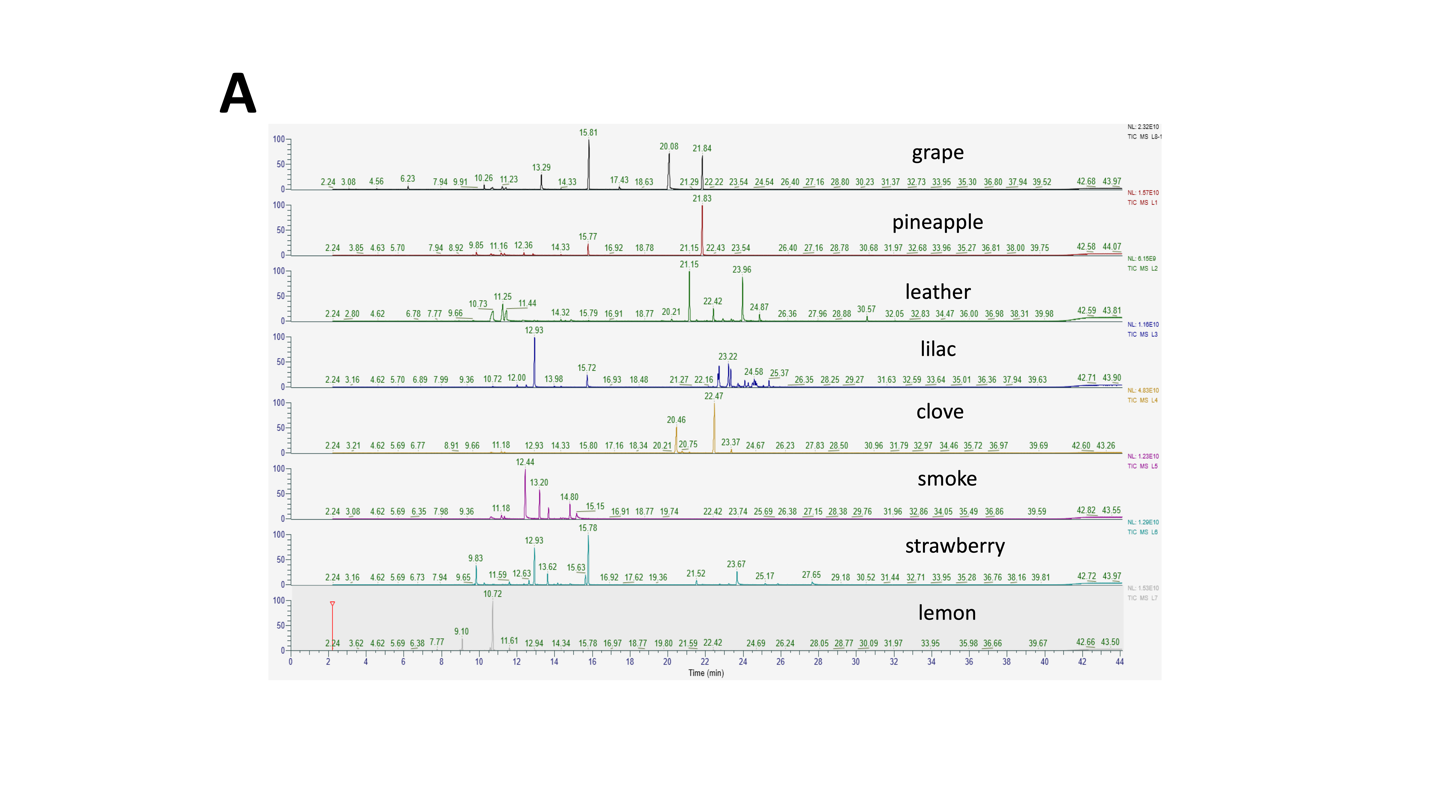

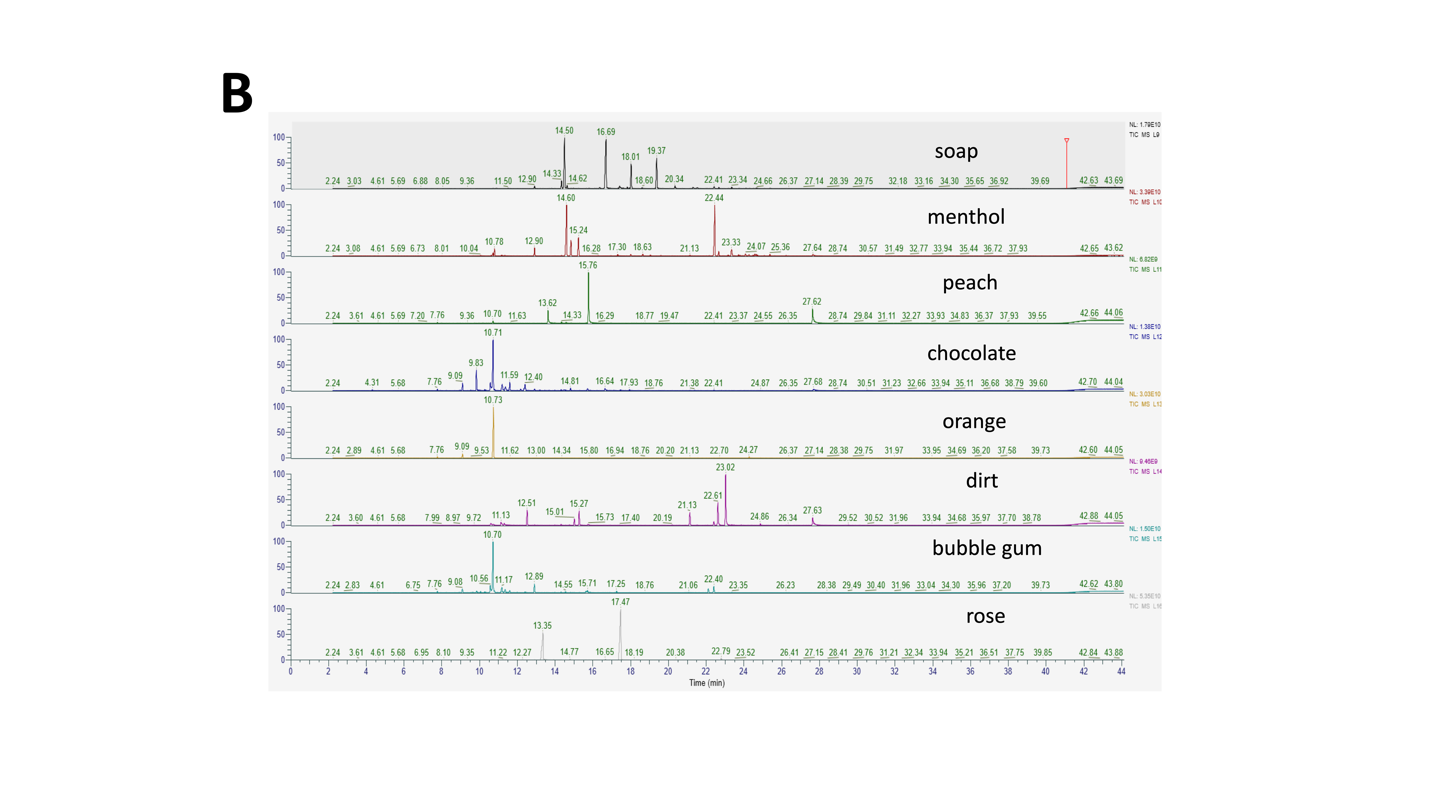

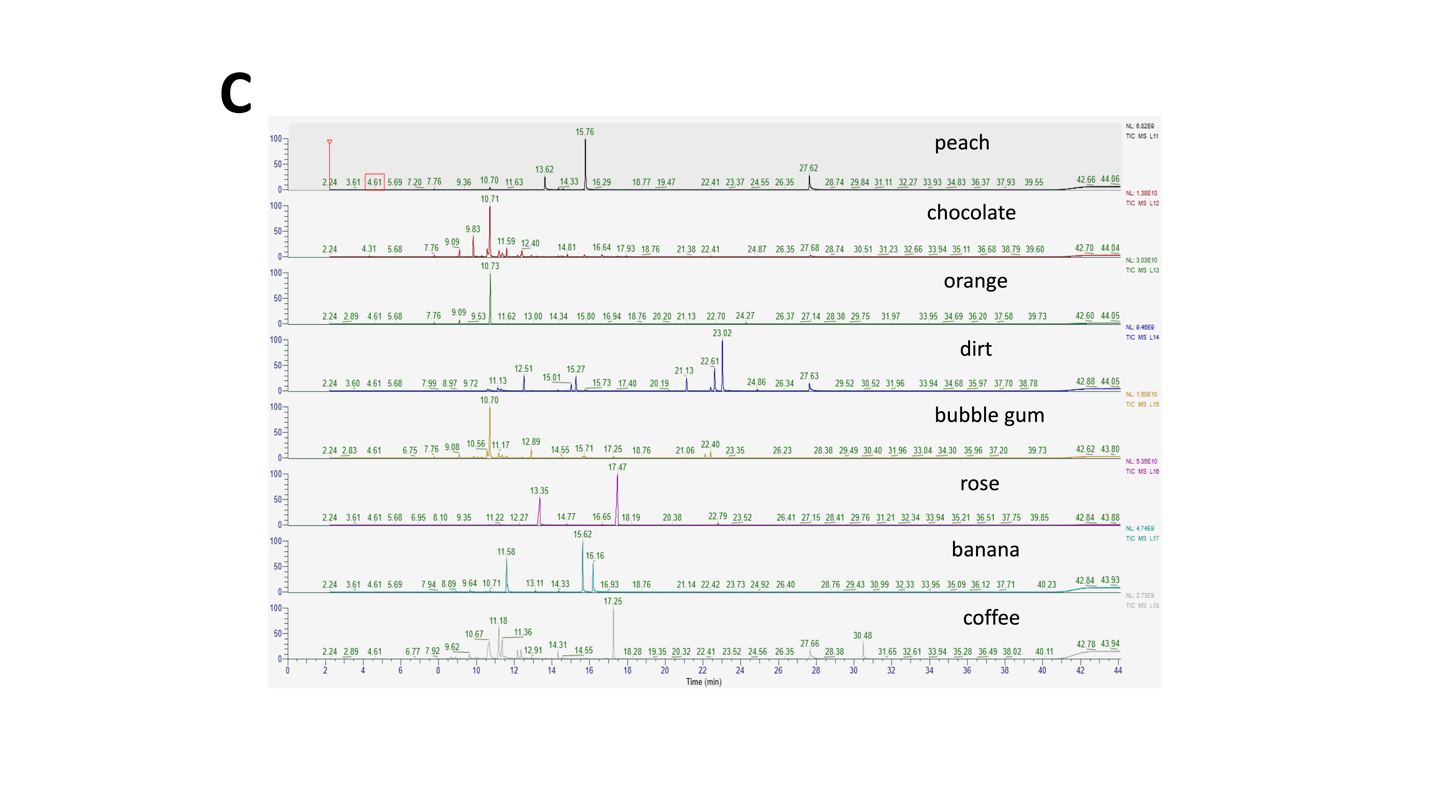

Extended Data Figure 2. Web-based application to collect responses. A) example of the intensity rating score collection. B) example of the OPID9 score collection. C) example of the POEM score collection. D) example of the OD10 score collection.

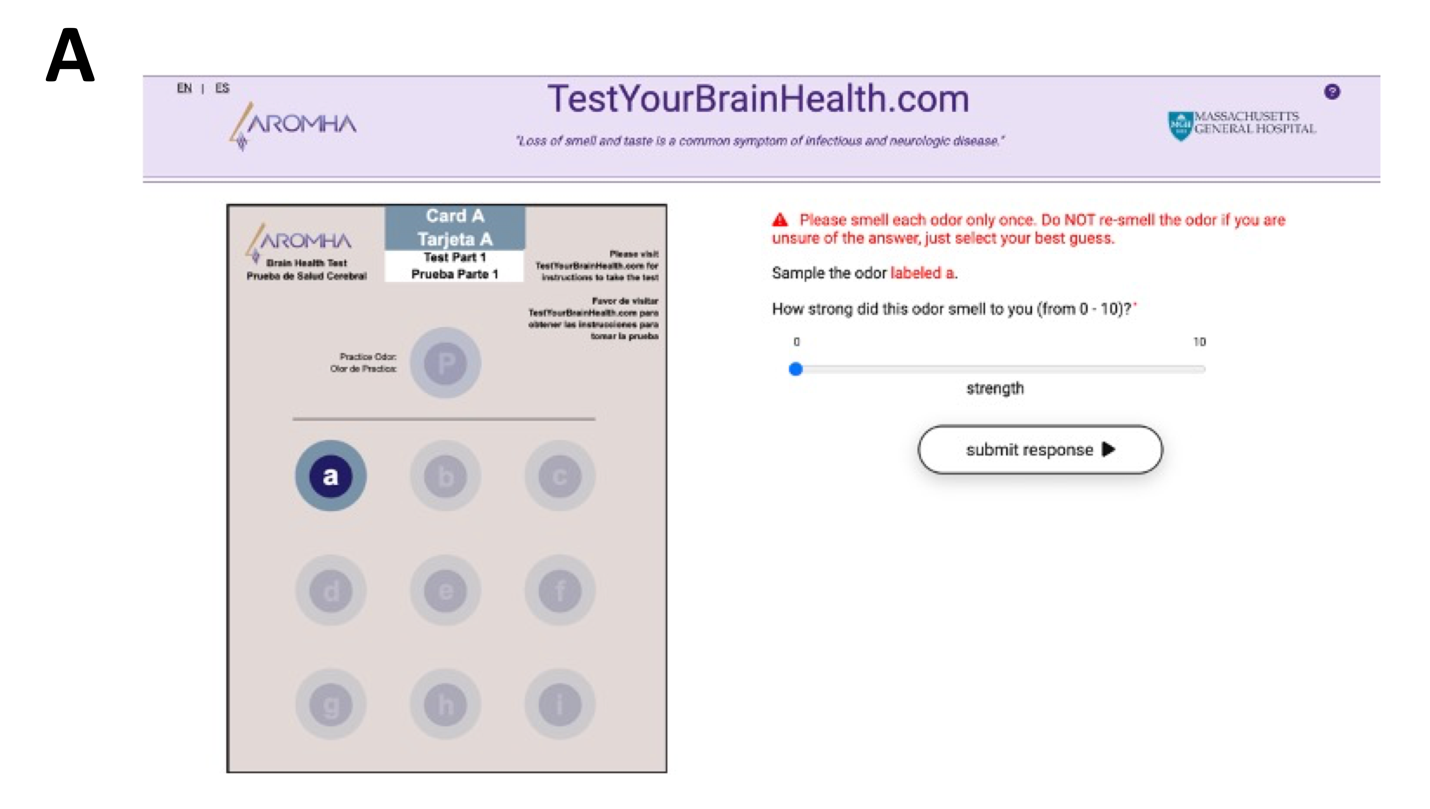

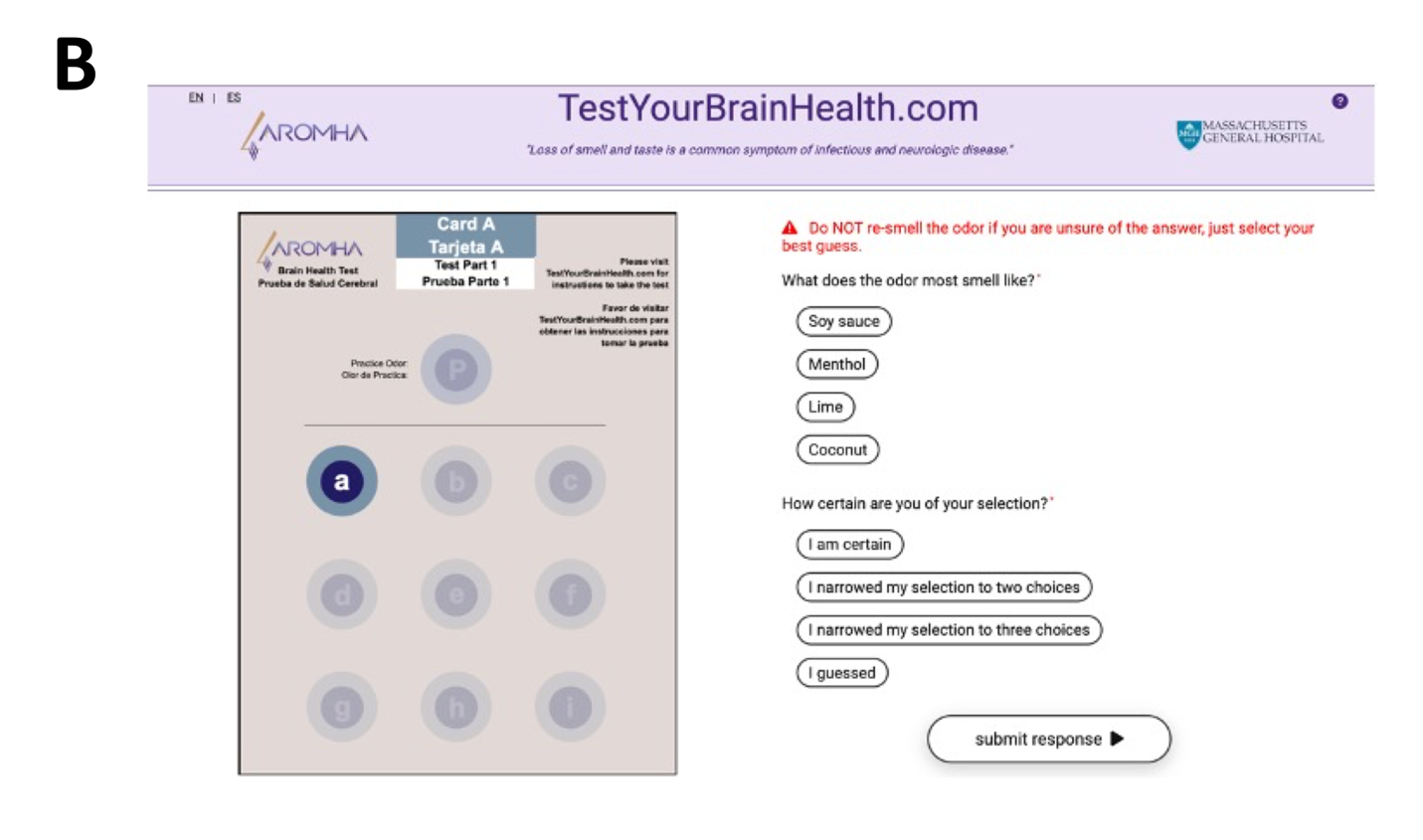

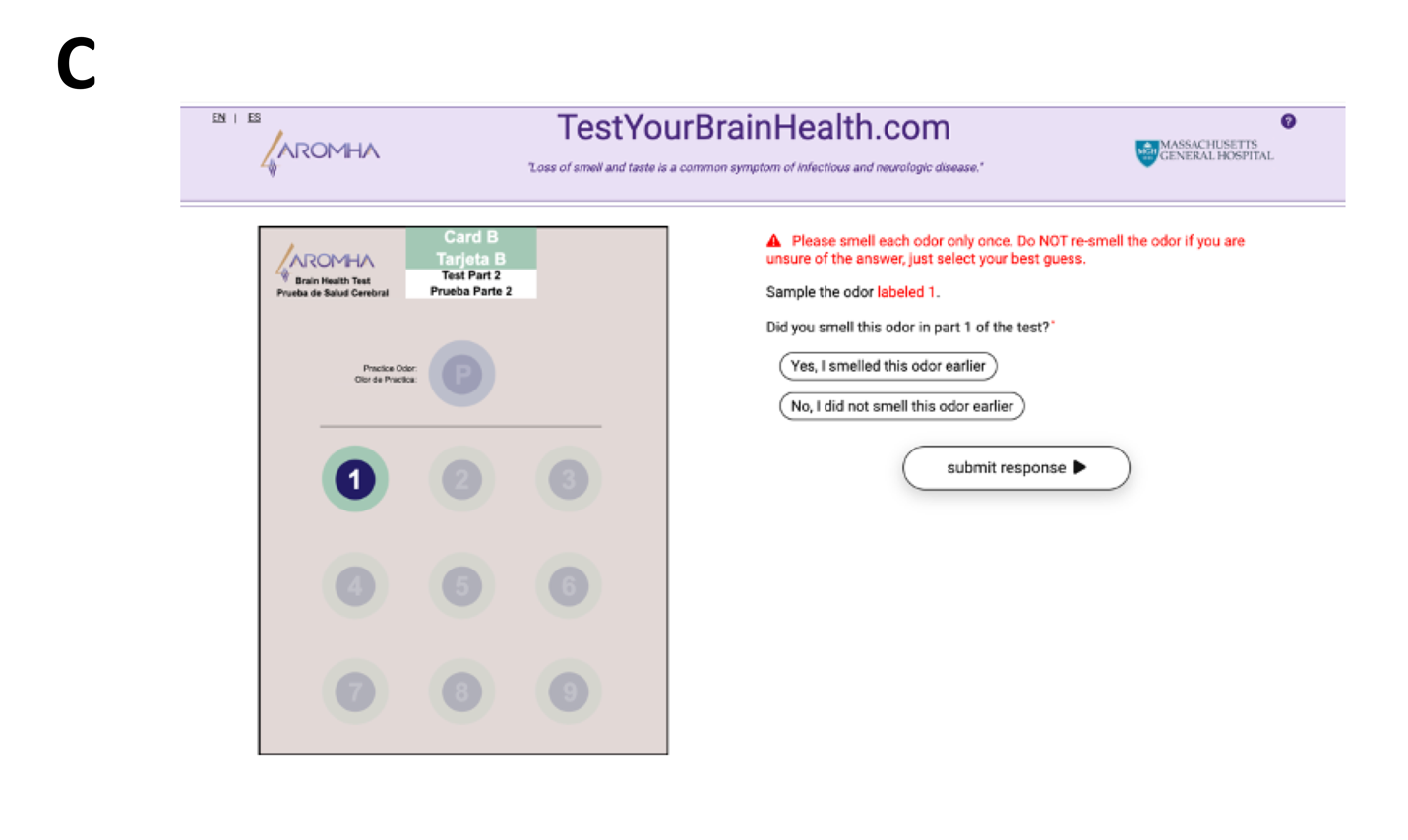

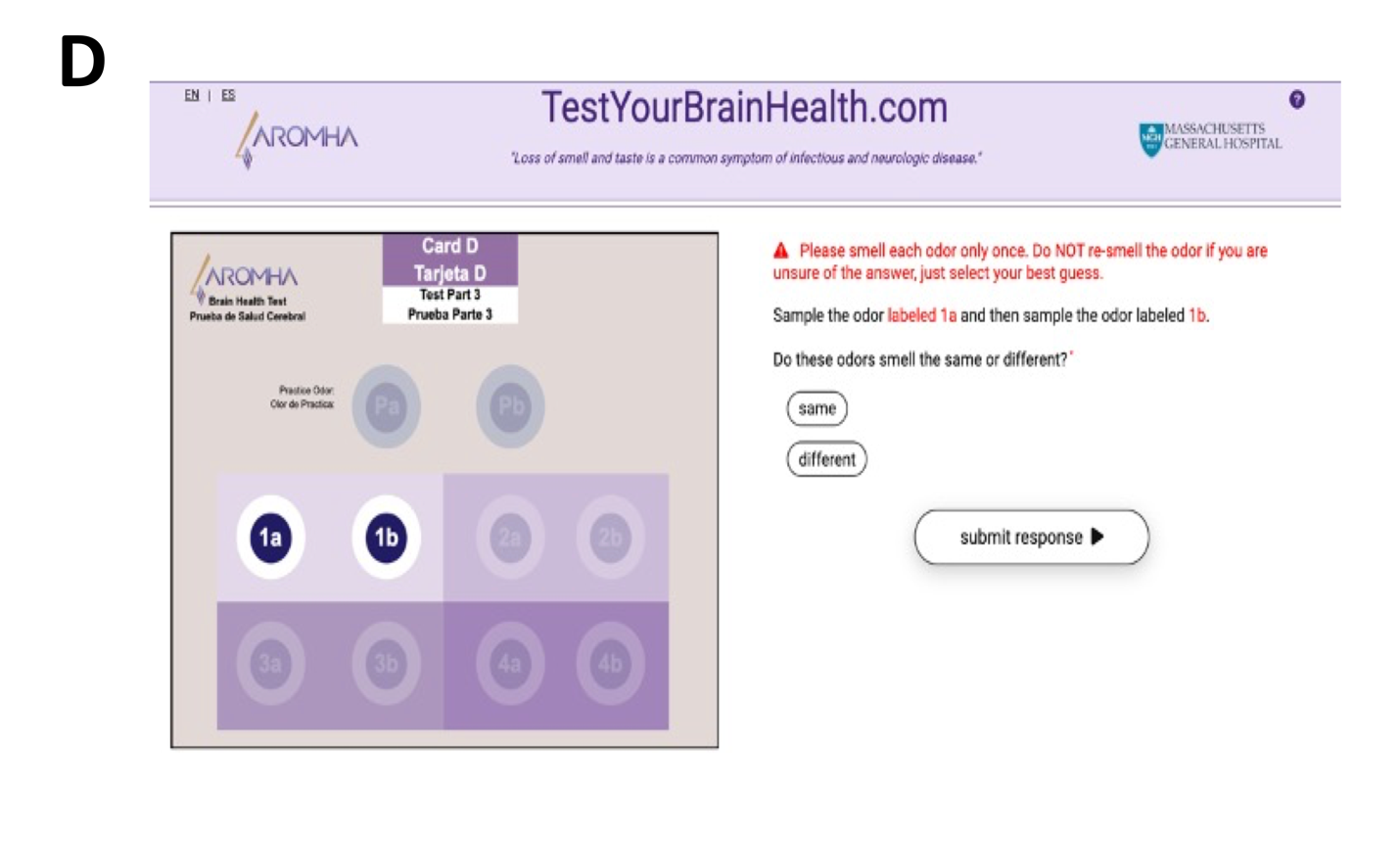
